# Tocilizumab versus baricitinib in hospitalized patients with severe COVID-19: an open label, randomized controlled trial

**DOI:** 10.1101/2022.06.13.22276211

**Authors:** Theodoros Karampitsakos, Ourania Papaioannou, Panagiota Tsiri, Matthaios Katsaras, Andreas Katsimpris, Andreas P. Kalogeropoulos, Elli Malakounidou, Eirini Zarkadi, Georgios Tsirikos, Vasiliki Georgiopoulou, Vasilina Sotiropoulou, Electra Koulousousa, Charikleia Chourpiliadi, Apostolos Matsioulas, Maria Lagadinou, Fotios Sampsonas, Karolina Akinosoglou, Markos Marangos, Argyris Tzouvelekis

## Abstract

**Background:** Tocilizumab and baricitinib have proven efficacy in COVID-19. There were no randomized-controlled trials comparing these compounds in patients with COVID-19.

**Materials/Patients and Methods:** In this open label, randomized controlled trial, we assigned 251 patients with COVID-19 and PaO_2_/FiO_2_<200 to receive either tocilizumab (n=126) or baricitinib (n=125) plus standard of care. To determine whether baricitinib was non-inferior to tocilizumab, we assessed if the upper boundary of the two-sided 95% confidence interval of the hazard ratio did not exceed 1.50. The primary outcome was mechanical ventilation or death by day 28. Secondary outcomes included time to hospital discharge by day 28 and change in WHO progression scale at day 10.

**Results:** Baricitinib was non-inferior to tocilizumab for the primary composite outcome of mechanical ventilation or death by day 28 (HR 0.83, 95% CI: 0.56 to 1.21, p=0.001 for non-inferiority). Baricitinib was non-inferior to tocilizumab for the time to hospital discharge within 28 days (discharged alive-tocilizumab: 52.4% vs baricitinib: 58.4%; HR 0.85, (95% CI: 0.61 to 1.18), p<0.001 for non-inferiority). There was no significant difference between baricitinib and tocilizumab arm in the change in WHO scale at day 10 [0.0 (95% CI: 0.0 to 0.0) vs 0.0 (95% CI: 0.0 to 1.0), p=0.83].

**Conclusion:** Baricitinib was non-inferior to tocilizumab with regards to the composite outcome of mechanical ventilation or death by day 28 and the time to discharge by day 28 in patients with severe COVID-19. Cost-effectiveness should be taken into account to avoid a dramatic upswing in health system budgets.

## Introduction

The emergence of 2019 coronavirus disease (COVID-19) is causing a growing global public health crisis^1^. Despite major advances in the prevention and treatment of COVID-19, a substantial proportion of infected individuals still experiences severe respiratory failure^2-5^. Emerging data suggest that hypoxic respiratory failure may be due in part to dysregulated inflammatory responses^6,7^. Thus, despite the benefits of anti-viral compounds such as remdesivir, extensive research efforts aimed to test the efficacy of compounds able to mitigate the immune response and prevent hyperinflammatory state. Corticosteroids had been consistently associated with improved survival across multiple studies. However, mortality of hospitalized patients with COVID-19 was still high despite corticosteroids implementation and the need for agents targeting deregulated immune responses was amenable^2,6,8^.

The IL-6R antagonist, tocilizumab, led to survival benefit in patients with COVID-19 irrespective of levels of inflammatory markers such as C reactive protein and ferritin^9-11^. Most recently, the oral selective Janus kinase 1/2 inhibitor, baricitinib, has been associated with reduced mortality in hospitalized patients with COVID-19^12-14^. Both compounds have been originally introduced as therapeutic modalities for several autoimmune diseases through downregulation of IL-6 levels, a cytokine implicated in endothelial and vascular dysfunction induced by SARS-CoV-2^11,14-17.^ Further investigation showed that baricitinib inhibited the intracellular signaling pathway of cytokines known to be induced by SARS-CoV-2, such as IL-2, IL-10, granulocyte– macrophage colony-stimulating factor and interferon-γ. Importantly, the biochemical inhibitory properties of baricitinib on human Numb-associated kinases (BIKE, AAK1 and GAK) implicated in SARS-CoV-2 viral propagation were also confirmed ^18^.

On the basis of the above, U.S. Food and Drug Administration issued an emergency use authorization for the use of both compounds in hospitalized patients with severe COVID-19. To this end, there is a paucity of high quality, randomized controlled trials comparing tocilizumab and baricitinib in patients with COVID-19. Limited data arise from retrospective, observational studies^19,20^. Given that the cost of baricitinib is considerably lower compared to tocilizumab, we aimed to conduct the first head-to-head randomized controlled trial and investigate whether baricitinib was non-inferior to tocilizumab in patients with severe COVID-19.

## Methods

### Trial design and oversight

We conducted an investigator-initiated, open-label, randomized-controlled trial enrolling consecutive patients with positive polymerase chain reaction test for SARS-CoV-2 admitted to our hospital between 20/10/2021 and 7/5/2022. Trial sites were three different COVID-19 departments of University Hospital of Patras, Greece. Each patient or the patient’s legally authorized representative provided written informed consent. The trial was conducted in accordance with the International Conference on Harmonisation E6 guidelines for Good Clinical Practice, the Declaration of Helsinki and the local regulations. Our study was approved by our Institutional Review Board and the Local Ethics Committee of University Hospital of Patras, Greece (Protocol Number: 26651/18-10-21). This was a registered clinical trial **(NCT05082714, https://clinicaltrials.gov/ct2/show/NCT05082714)**. The study design is provided in **Figure 1**.

**Figure 1.**
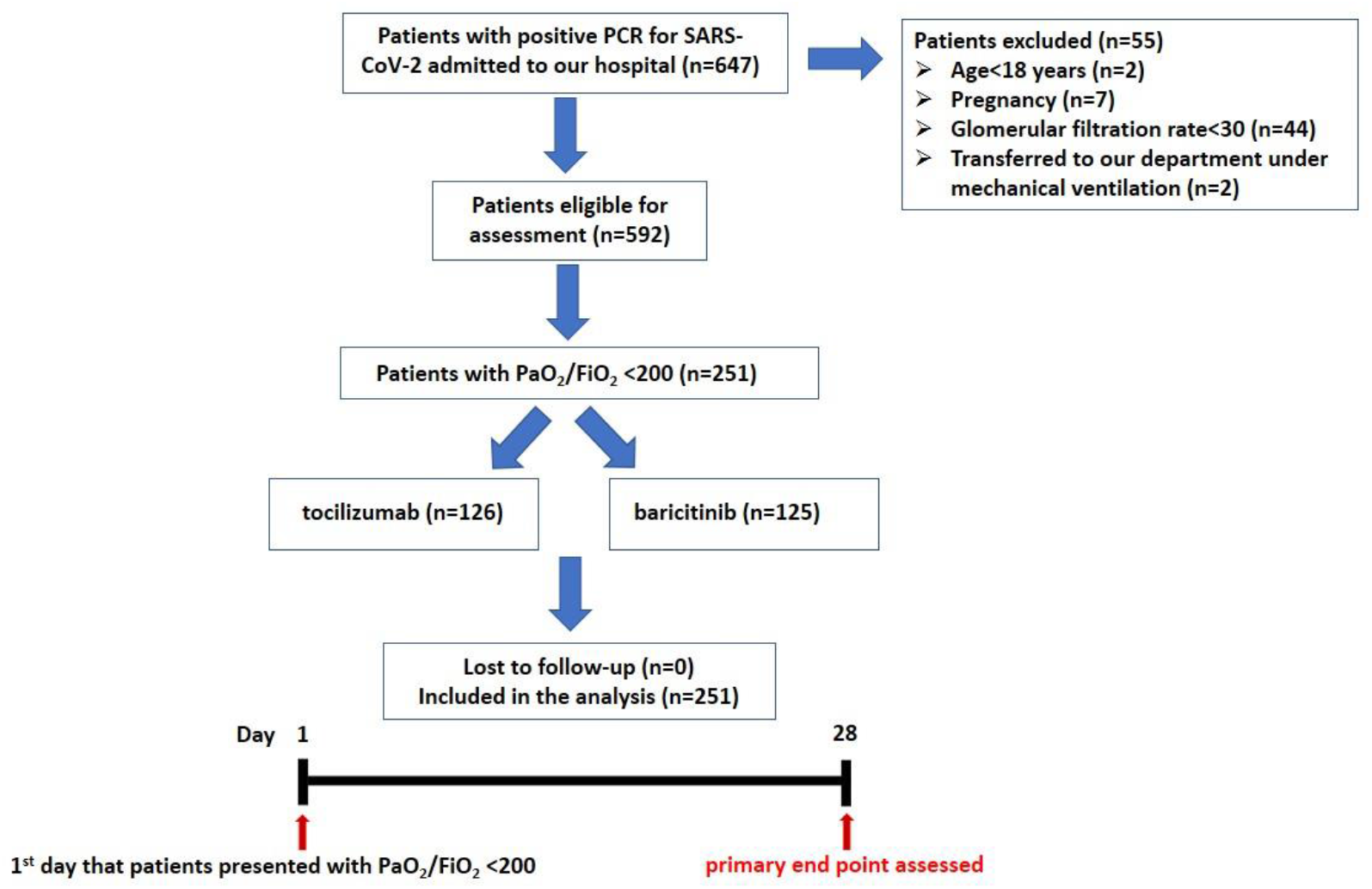
Schematic representation of the study design.

### Participants

Patients aged 18 years or older that presented with PaO_2_/FiO_2_ <200 at any time during their hospitalization were included in the analysis. Exclusion criteria were: age<18 years, pregnancy, estimated glomerular filtration rate<30 mL/min/1.73m^2^ and mechanical ventilation prior to patients’ transfer to our Hospital.

### Randomization and interventions

Day 1 was considered the first day when a patient reached a PaO_2_/FiO_2_ <200. Patients were randomly assigned through a random sequence generator (derived from http://www.randomization.com) to tocilizumab or baricitinib on 1:1 ratio. Treatment with tocilizumab or baricitinib started on Day 1 at the time point that PaO2/FiO_2_<200 was recorded. Tocilizumab was administered as a single infusion intravenously at 8mg/kg. Baricitinib was administered at a dose of 4 mg/day (given daily for up to 14 days or until discharge from hospital, whichever occurred first) or at a dose of 2 mg/day in case of estimated glomerular filtration rate of 30 to less than 60 mL/min/1.73m^2^. All patients included in the analysis received standard of care including : 1) dexamethasone at a dose of 6mg/day and the dose did not change in any patient, 2) remdesivir 200 mg loading dose and then 100 mg per day for 5 days, 3) anticoagulants (low molecular weight heparin at prophylactic dose). Standard of care regimens in a minority of cases also included: antibiotic compounds and vasopressor support that were administered upon clinicians’ judgment. Standard of care did not include convalescent plasma, nintedanib and pirfenidone. The study design is summarized in **Figure 1**.

### Outcome measures

The primary end point was mortality or mechanical ventilation by day 28. Our secondary outcomes were 1) time to discharge by day 28 and 2) disease progression as indicated by change in WHO clinical progression scale at day 10[ΔWHO scale (day 10 - day 1)].

### Data collection

We recorded PaO_2_/FiO_2_ of each patient, as well as demographics, comorbidities, laboratory parameters, WHO clinical progression scale, time to discharge and time to event (mortality or mechanical ventilation). WHO clinical progression scale was used as a measure of illness severity across a range from 0 (not infected) to 10 (dead) with data elements being easily obtainable from clinical records^21^.

We recorded patients that presented with platelet count above 450 × 10^9^/L, 5-fold increased creatine phosphokinase greater than the reference value and 3-fold increased transaminases compared to reference value. The incidence of other adverse events such as lobar consolidation, major bleeding, cardiac event and septic shock was also recorded. These events were evaluated according to the National Cancer Institute Common Terminology Criteria for Adverse Events, version 5.0.

### Sample size

We initially planned for a total sample size of 164 patients. However, on the basis of fast recruiting, we submitted a protocol modification and increased the total sample size to 251 patients without changing outcome measures. The final sample size was estimated under the following assumptions: (1) a randomization ratio of 1:1 for tocilizumab and baricitinib; (2) a two-sided type I error a of 0.05; (3) Hazard Ratio (HR) θ: 1; (4) HR θo: 1.5 (the upper boundary of the two-sided 95% confidence interval of the HR for the risk of the primary composite endpoint not exceeding 1.50); (5) power of 80%; (6) based on our previous records, 60% of patients receiving standard of care required mechanical ventilation or died within 28 days after PaO_2_/FiO_2_ <200 was reached.

### Statistical Analysis

The primary outcome was presented using the Kaplan–Meier method and cumulative incidence curves were compared between the two groups. We assessed the primary non-inferiority hypothesis by investigating if the upper boundary of the two-sided 95% confidence interval of the HR for the risk of the primary composite endpoint did not exceed 1.50. Time-to-discharge was presented with the use of the Kaplan–Meier approach. We assigned the ‘worst outcome’ for individuals who died before day 28 and therefore these patients were managed as those with the longest hospital stay (more than 28 days)^22^. Non-inferiority was assessed for the secondary outcome of time to hospital discharge. Mann-Whitney U test was used for the detection of differences in ΔWHO scale between the two arms. p-values < 0.05 were considered statistically significant.

## Results

### Patients

Between 18/10/2021 and 7/5/2021, 251 patients were randomly allocated to receive either tocilizumab plus standard of care (n=126) or baricitinib plus standard of care (n=125). No participant was lost to follow-up. All patients completed the trial and were included in the analysis (n=251/251, 100%). Baseline demographics and disease characteristics were similar between study groups **(Table 1)**. Median age (interquartile range) was 72.0 (62.0 to 83.0) and 73.0 (61.0 to 83.0) years in the tocilizumab and baricitinib group, respectively. The majority of patients were male in both groups [tocilizumab: n=74, (58.7%), baricitinib: n=74, (59.2%)]. Arterial hypertension was the most common comorbidity both in the tocilizumab (n=67, 53.2%) and baricitinib arm (n=67, 53.6%) **(Table 1)**.

**Table 1.**
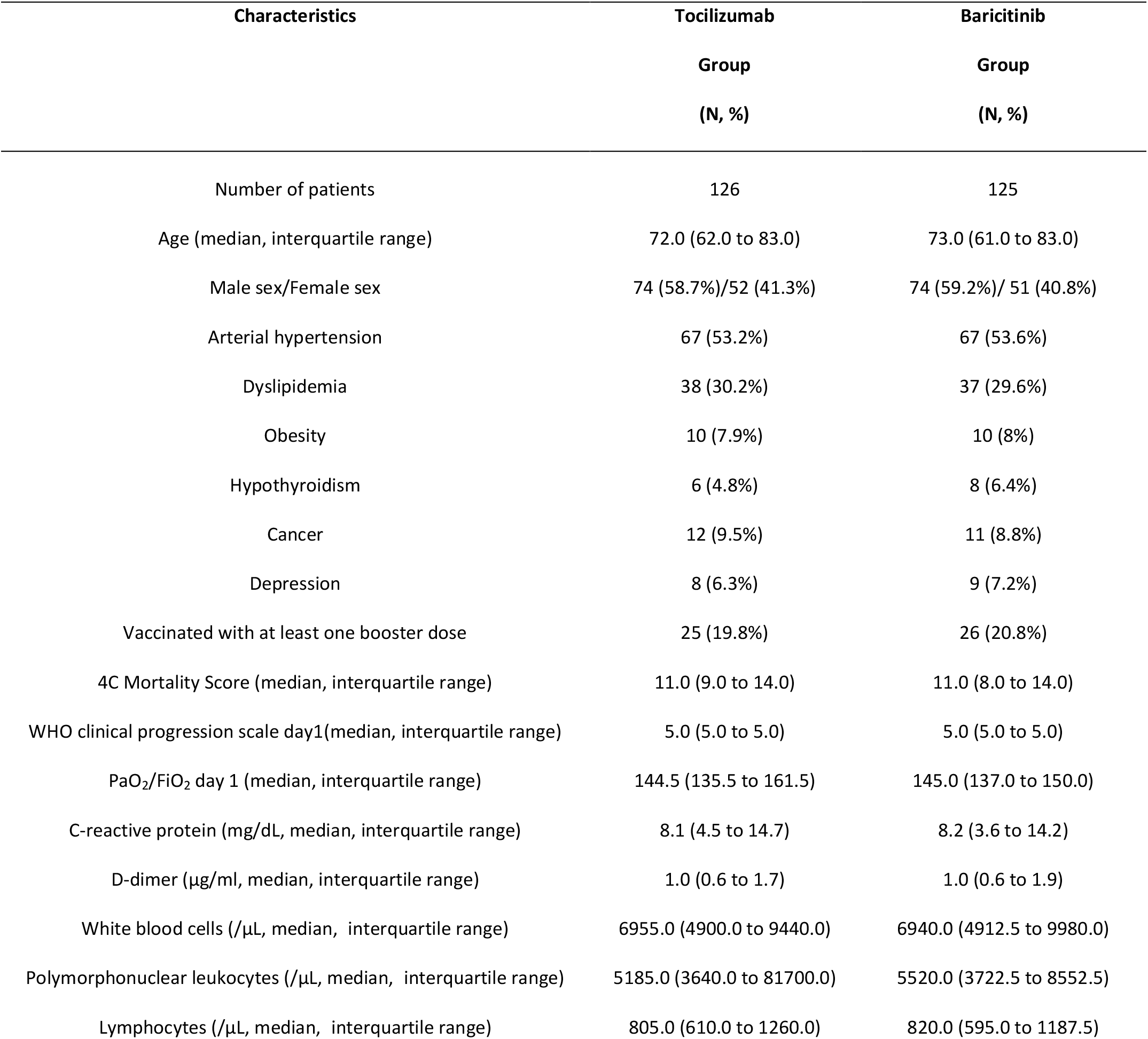
Characteristics of patients at baseline.

### Primary outcome

Baricitinib was non-inferior to tocilizumab for the primary composite outcome of mechanical ventilation or death by day 28 (HR 0.83, 95% CI: 0.56 to 1.21, p=0.001 for non-inferiority), **(Figure 2A)**. Mechanical ventilation or death by day 28 occurred in 39.2% (n=49) and 44.4% (n=56) of patients in the baricitinib and tocilizumab arm, respectively.

**Figure 2.**
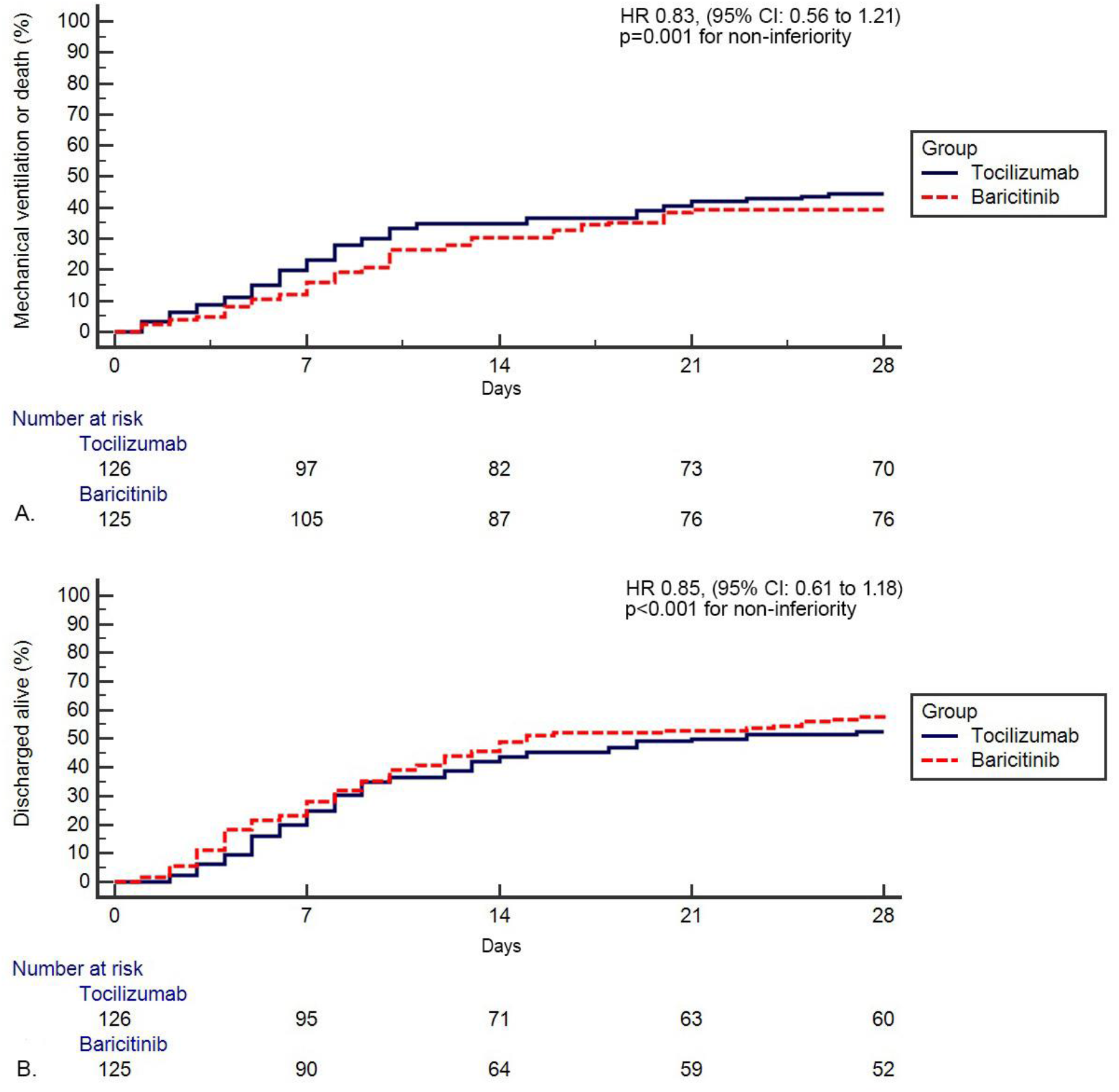
Kaplan-Meier curve showing the effect of allocation to baricitinib or tocilizumab on mechanical ventilation or death by day 28 **(Panel A)** and on discharge from hospital within 28 days of randomization **(Panel B)**.

### Secondary outcomes

Baricitinib was non-inferior to tocilizumab for the time to hospital discharge within 28 days [HR 0.85, (95% CI: 0.61 to 1.18), p<0.001 for non-inferiority], **(Figure 2B)**. Seventy three patients (n=73, 58.4%) in the baricitinib group and 66 (52.4%) in the tocilizumab group had been discharged alive within the 28-day period. Among patients that were discharged alive within the 28-day period, median days of hospitalization were 8.0 (95% CI: 6.0 to 10.0) and 8.0 (95% CI: 6.6 to 9.0) in the baricitinib and tocilizumab group, respectively. There was no significant difference between baricitinib and tocilizumab arm in the risk of disease progression by day 10, as assessed by change in WHO scale at day 10 [ΔWHO scale (day 10 - day 1) - baricitinib: 0.0 (95% CI: 0.0 to 0.0) vs tocilizumab: 0.0 (95% CI: 0.0 to 1.0), p=0.83], **(Figure 3A)**.

**Figure 3.**
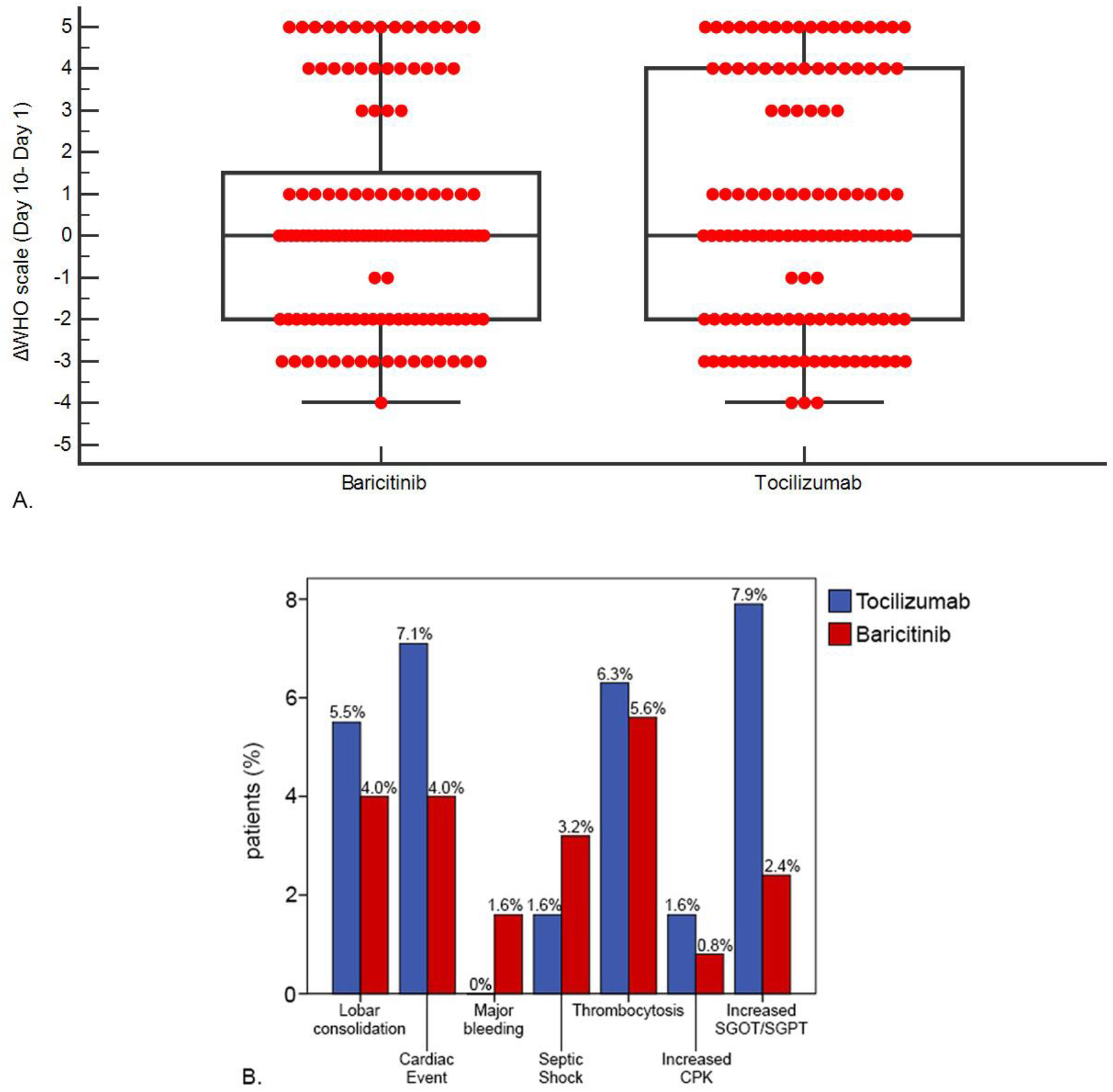
Mann–Whitney U test investigating difference in change in WHO scale at day 10 [ΔWHO scale (day 10 - day 1) **(Panel A)**. Adverse events of patients allocated to tocilizumab and baricitinib **(Panel B)**.

### Adverse events

There were 33 (26.2%) and 25 (20%) patients with at least one treatment-related adverse event in the tocilizumab and baricitinib arm, respectively **(Table 2, Figure 3B)**. There was a statistically significant difference in the number of patients presenting with 3-fold increased transaminases between tocilizumab (n=10, 7.9%) and baricitinib arm (n=3, 2.4%), (p=0.04). A similar distribution of other treatment-related adverse events such as lobar consolidation, thrombocytosis and increased creatine phosphokinase was observed.

**Table 2.** Adverse events by day 28.

| Adverse events | Tocilizumab<br>Group<br>(N=126) | Baricitinib<br>Group<br>(N=125) | p<br>value |
| --- | --- | --- | --- |
| Lobar consolidation | 7 (5.5%) | 5 (4.0%) | 0.57 |
| Cardiac Event | 9 (7.1%) | 5 (4.0%) | 0.28 |
| Major bleeding | 0 (0.0%) | 2 (1.6%) | 0.15 |
| Septic shock | 2 (1.6%) | 4 (3.2%) | 0.41 |
| Thrombocytosis | 8 (6.3%) | 7 (5.6%) | 0.82 |
| Increased CPK 5 times greater than the upper<br>reference value | 2 (1.6%) | 1 (0.8%) | 0.56 |
| Increased SGOT/SGPT 3 times greater than the<br>upper reference value | 10 (7.9%) | 3 (2.4%) | 0.04 |

**Abbreviations:** CPK: Creatine phosphokinase, SGOT: Serum glutamic-oxaloacetic transaminase, SGPT: serum glutamic-pyruvic transaminase

## Discussion

This is the first head-to-head randomized-controlled trial showing that baricitinib was non-inferior to tocilizumab with respect to the primary composite outcome of mechanical ventilation or death by day 28 in hospitalized patients with severe COVID-19. Baricitinib was non-inferior to tocilizumab for the time to hospital discharge within 28 days. There was no significant difference between baricitinib and tocilizumab in the change of WHO progression scale at day 10. The aforementioned findings are of paramount importance, given that we investigated a relatively homogenized cohort of patients with severe COVID-19.

In particular, our study exhibited a number of important attributes that should be presented upfront. First, we enrolled a relatively homogenized group of patients with severe disease based on physiological indices (PaO_2_/FiO_2_ <200) under standardized usual care including remdesivir and corticosteroids that received baricitinib or tocilizumab precisely at the time of clinical and physiological deterioration, as assessed by PaO_2_/FiO_2_ <200. Second, all patients were managed in the same hospital from the same clinicians under a common therapeutic algorithm. Thus, outcomes were not affected by heterogeneous approaches. Third, our results could maximize cost-effectiveness in the management of COVID-19 and limit the dramatic upswing in health system budgets^23^. To this end, there is a paucity of biologically enriched studies able to demonstrate patients who are more likely to experience benefit from tocilizumab than baricitinib. On the other hand, the cost of baricitinib is substantially lower than the cost of tocilizumab. Moreover, baricitinib is administered per os in tablet form, has few drug–drug interactions and is excreted largely unchanged. Finally, baricitinib showed a favorable safety profile besides being non-inferior to tocilizumab in multiple efficacy outcomes of our study. Based on the above, adopting baricitinib as the first-line treatment for severe COVID-19 might be rational.

Our results are in line with previous high-quality trials in COVID-19. Baricitinib had shown the largest effect size on mortality of hospitalized patients with COVID-19 among immunomodulatory compounds used in other randomized controlled trials^13,24,25^. Baricitinib demonstrated benefit in addition to the use of corticosteroids alone^24^. A recent head-to-head trial between baricitinib and dexamethasone in COVID-19 showed similar rates of mechanical ventilation-free survival by day 29; yet, fewer adverse events were encountered in the baricitinib arm^26^. This might be explained by the fact that baricitinib has a shorter half-life than dexamethasone, exerts its anti-inflammatory role by acting on targeted critical pathways and thus biologic redundancy is minimized with less immunosuppression^13,27^. Further to these, baricitinib may exert synergistic effects with remdesivir antiviral properties^27,28^.

Contrary to the results of baricitinib trials in COVID-19, the first studies investigating the efficacy profile of tocilizumab in hospitalized patients with COVID-19 yielded contradictory results. In particular, a meta-analysis of the first eight randomized controlled trials of tocilizumab in COVID-19 did not show a survival benefit ^9,10,29-35.^ The RECOVERY trial, including an overall of 4116 patients with COVID-19, attenuated confusion by showing beneficial effects of tocilizumab with regards to hospitalization, need for mechanical ventilation and survival^9^. The above contradictory results led to a debate whether timing of administration influenced the efficacy of tocilizumab^36^. Our group had previously shown that tocilizumab was efficacious when applied at the time point that PaO_2_/FiO_2_ <200 was observed^11^. Therefore, we adopted that approach in this trial.

Baricitinib had shown consistent efficacy independent of disease severity^14,23,24^; yet, it has been suggested that the observed benefit might be more evident in patients with a WHO ordinal score of 5 (supplemental oxygen) or 6 (in need of high-flow oxygen or non-invasive ventilation)^13,37^. Patients presenting for the first time with PaO_2_/FiO_2_ <200 typically belong to a score of 5 or 6 in WHO ordinal scale. Importantly, enrolling patients precisely at the time point that PaO_2_/FiO_2_ falls below 200 renders our cohort considerably homogeneous with regards to respiratory status. Our ‘‘not too early, not too late’’ approach with regards to the time point of tocilizumab or baricitinib administration was based on three concepts. First, we aimed to avoid further immunomodulation at the early phases of SARS-CoV-2 replication. Second, patients with the most severe disease status might not benefit accordingly from these compounds, as inflammatory cascade might be too advanced to be reversible. The concept that deregulated immune responses arising from redundant immune signaling pathways can be even more detrimental than pathogens themselves is highly relevant in the context of COVID-19^28^; thus, timely intervention is crucial. The ideal time window to intervene might correlate with the time around clinical deterioration, as assessed by PaO_2_/FiO_2_ <200^11^. Third, administration of baricitinib or tocilizumab at the time point of clinical deterioration might limit their irrational, uncontrolled use, maximize cost-effectiveness and reduce immunocompromisation-related side effects.

Our trial has some limitations that should be treated cautiously. First, this is an open label randomized controlled trial. Despite that assessment of the primary efficacy outcome is objective, secondary outcomes including time to discharge and score in WHO progression scale are operator-dependent. Limitations associated with the use of WHO progression scale as a secondary outcome include lack of proportionality among categories, absence of an established minimum clinically important difference and heterogeneity in local clinical practice. However, management of our cohort from clinicians of the same hospital with a homogeneous approach represents the optimal way to maximize the value of the aforementioned secondary outcomes. Second, our sample size was moderate; yet, adequate to assess non-inferiority based on our pre-specified plan. Finally, given that this is the first head-to-head trial of these compounds in COVID-19, margin could not be derived from a previous well-designed trial. Thus, we chose the value that represented the median non-inferiority margin used in previous high-quality non-inferiority trials^38^. The power of our trial and type I error were 80% and 0.05 based on our assumptions, respectively. Despite that the quality of our study could be even higher with different assumptions, these assumptions are acceptable for non-inferiority randomized trials.

Collectively, this is the first head-to-head trial between baricitinib and tocilizumab in COVID-19. The results of this open label, randomized controlled trial demonstrated that baricitinib was non-inferior to tocilizumab in hospitalized patients with COVID-19 with regards to the composite outcome of mechanical ventilation or death by day 28 and the time to discharge by day 28 in patients with severe COVID-19. Given the absence of theragnostic biomarkers able to guide decision between tocilizumab and baricitinib for the treatment of a highly contagious disease of enormous economic burden, cost-effectiveness and optimal safety profile should be taken into consideration in the everyday clinical setting. Biologically enriched studies aiming to identify subgroups of patients that are more likely to benefit from targeted immunomodulatory compounds are eagerly awaited.

## Data Availability

Data are available upon request

## Data availability statement

data available upon request.

## Acknowledgment

Authors would like to thank the staff of our hospital.

